# Clinical validation and the evaluation of a colorimetric SARS-CoV-2 RT-LAMP assay identify its robustness against RT-PCR

**DOI:** 10.1101/2022.03.21.22271747

**Authors:** M. Erdem, A. Andac-Ozketen, AC. Ozketen, G. Karahan, A. Tozluyurt, F. Palaz, A. Alp, S. Unal

## Abstract

The novel coronavirus has infected millions of people all around the world and has posed a great risk to global health. Rapid and accurate tests are needed to take early precautions and control the disease. The most routinely used method is real time polymerase chain reaction (RT-PCR) which stands as the gold standard in the detection of SARS-COV-2 viral RNA. However, robust assays as accurate as RT-PCR have been developed for rapid diagnosis and efficient control of the spread of the disease. Reverse transcriptase loop-mediated isothermal amplification (RT-LAMP) is one of the time-saving, accurate and cost-effective alternative methods to RT-PCR. In this study, we study the improved RT-LAMP colorimetric assay (N-Fact) to detect SARS-COV-2 viral RNA within 30 minutes using a primer sets special to N gene. Moreover, RT-LAMP colorimetric assay is subjected to authorized clinical studies to test its ability to detect COVID-19 in its early phases. The results reveal RT-LAMP colorimetric assay is an efficient, robust, and rapid assay to be used as in vitro diagnostic tool display competitiveness compared to RT-PCR.

## INTRODUCTION

In December 2019, a novel coronavirus (SARS-CoV-2) that causes coronavirus disease 2019 (COVID-19) emerged from its epicenter in Wuhan province in China [1]. The rapid spread of this disease has threatened global public health seriously and it has a huge impact on the global economy. In the wake of the rapid spread of the coronavirus, there has been an urgent need for rapid and sensitive detection systems. Hence, suspected cases are effectively identified, patients are rapidly screened and virus surveillance is conducted for isolation strategies [2].

The current standard method which is used to detect SARS-CoV-2 is RT-PCR [3, 4]. Regardless of its high sensitivity and specificity, this method has some drawbacks as it requires complex and expensive equipment, extensive training for users, and multiple hours to obtain the result. These weaknesses limit the screening capacity of the RT-PCR technique and it falls behind the rapid growing SARS-CoV-2 cases [5, 6]. As a result, more efficient methods are in high demand for the detection of COVID-19 to catch its growing pace. Recently, loop-mediated reverse transcription loop-mediated isothermal amplification (RT-LAMP) has been used as an alternative to the RT-PCR method. RT-LAMP has many advantages over RT-PCR such as requiring a simple instrument (e.g., heating block) and a constant temperature for amplification, and gives result in a short period of time [7]. Moreover, the results can be visualized by the naked eye with the presence of a colorimetric pH indicator. Since having the same sensitivity and specificity as RT-PCR, the RT-LAMP method is a more effective choice for high-throughput and low-cost detection of SARS-CoV-2 [8, 9].

In this research, the RT-LAMP method was used to detect SARS-COV-2 in 30 min at a constant 65 °C and positive samples were detected by the color change from fuchsia to yellow. Phenol red was used as a dye indicator in which as the reaction proceeds, the pH of the reaction solution gets lower and causes a color change. To validate the sensitivity and specificity of the RT-LAMP assay, the results were compared with RT-PCR results. The clinical studies were conducted on 300 naso/oropharyngeal swab specimens collected from COVID-19 suspects. We tested and compared RT-LAMP and RT-PCR results on the samples to characterize efficiency of the RT-LAMP colorimetric assay in Hacettepe University Faculty of Medicine. The study reveal RT-LAMP is indeed a reliable assay exceeding RT-PCR in terms of time interval used, cost effectiveness and robustness.

## MATERIALS AND METHODS

### Clinical Sample Handling

Samples were collected as nasopharyngeal swabs in virus inactivation medium (vNAT; Bioeksen R&D Technologies Ltd, Turkey). vNAT buffer extracted and preserved viral nucleic acids in respiratory tract samples. The sample collection occurred as part of the routine operation of Hacettepe University Faculty of Medicine and each sample was used both RT-LAMP and RT-PCR testing.

### RT-PCR

RT-PCR was performed by using Bio-Speedy SARS-CoV-2 RT-PCR kit (Bioeksen R&D Technologies Ltd, Turkey). Detection with the kit was achieved via rapid nucleic acid extraction from respiratory tract samples followed by multiplex RT-PCR targeting the SARS-CoV-2 specific *ORF1ab* gene and human *RNase P* gene and mRNA in CFX96 real time PCR instrument (Bio-Rad, USA). The oligonucleotide set targeting human *RNase P* gene and mRNA functioned as a control of the sampling, nucleic acid extraction and inhibition.

### RT-LAMP Primer Design

RT-LAMP assay has six primers which are two inner primers (FIP and BIP), and two outer primers (F3 and B3) and two loop primers (Forward loop primer; LF, and backward loop primer; LB) (11,12). RT-LAMP primers were designed by Primer Explorer (https://primerexplorer.jp/e/) for the N-gene of SARS-CoV-2. For positive control, N-gene sequence was obtained from National Center of Biotechnology Information (NCBI) with GenBank accession number MN908947.3 and genomic positions between 27894..28259 of Wuhan-Hu-1 genome (NCBI Reference Sequence: NC_045512) was used which was cloned in pGEM®-T Easy vector.

### RT-LAMP Assay

First, the swab samples in virus inactivation medium (vNAT) were lysed with N-fast lysis buffer at 95 °C for 5 min in which they were mixed at 1:1 portion. Then, the RT-LAMP mix was prepared. For one reaction the total volume is 25 µL: 18.5 µL mastermix, 2.5 µL primer mix (10X) (16 µM FIP/BIP, 2 µM F3/B3, 4 µM LF/LB), 2.5 µL A1 (10X), 0.5 µL A2 (50X) and 1 µL was taken from lysed swab sample (Table 1). For positive control, N-gene plasmid (0.5 ng) was mixed with vNAT and quick lysis solution in 2:1:1 proportion. For the negative control 1:1 portion lysis buffer and vNAT were used. The reaction mixture was incubated at 65 °C for 30 min and at the end of the reaction, the color difference between positive and negative samples were analyzed with naked eye. The positive samples were seen as yellow color and the negative ones were purple.

**Table 1.** n-Fast kit components and reaction volumes

| Components | Volume (μL) | Final Concentration |
| --- | --- | --- |
| Master mix | 18.5 | - |
| Primer mix | 2.5 | 1X |
| A1 Reagent | 2.5 | 1X |
| A2 Reagent | 0.5 | 1X |
| Sample | 1 | - |
| Total | 25 |  |

### Testing Clinical RNA Samples with the RT-LAMP Assay

For evaluating the n-Fast Rt-LAMP kit, it was applied to Turkish Republic Ministry of Health, Turkish Drugs and Medical Devices Agency with a number of E.471441. The volunteers of 300 applying for the test at Hacettepe University Hospital with their consent occurring from November 3 through November 24, 2020 were included in the study. Of the volunteers, 150 were selected from those who did not show symptoms of COVID-19, and 150 from those who showed symptoms. Combined nose-throat swabs were taken from the volunteers once and the samples were transferred to Hacettepe University Central Laboratory, Department of Molecular Microbiology under appropriate conditions. The samples were labeled by recording the date, presence of symptoms, and the anonymous number given to the patient. All the samples were tested simultaneously with both the n-Fast kit we developed for the diagnosis of COVID-19 and the RT-PCR method used in routine practice. Analyzing the consistency of the results from both analytic tests were later compared and the inconsistent test results were further investigated considering the clinical and epidemiological findings, laboratory results, and radiological evaluations of the volunteers.

## RESULTS

### Sensitivity of the RT-LAMP assay in detecting SARS-CoV-2 using N gene

Colorimetric RT-LAMP assay was developed to detect the nucleocapsid (N) gene of SARS-COV-2. To evaluate the sensitivity of RT-LAMP assay, serial dilutions of 5*10^−3^ ng DNA from 1:10^2^ to 1:10^9^ were prepared from the N-gene plasmid. Because of the preparation of the N-gene plasmid in 2:1:1 proportion with vNAT and lysis buffer, the final concentrations of the diluted N-gene plasmid solutions became 5* 10^−4^ to 5*10^−9^ ng/µL. The color change from purple to yellow in 30 minutes was observed up to 106 dilution which is equal to 5*10^−6^ ng (Fig. 1). In addition, number of copies of the cloned N-gene was quantified as the stated equation: Copies/μL = concentration of plasmid (g/μL)/[(plasmid length × 660) × (6.022 × 10^23^)] and our RT-LAMP assay was detected the N-gene up to nearly 60 copies/µL (Table 2) [10].

**Table 2.** Comparison the sensitivity of RT-LAMP and RT-PCR results.

| Dilution | RT-LAMP |  | RT-PCR |  | Ct value |
| --- | --- | --- | --- | --- | --- |
|  | Result | DNA load(copies/<br>rxn) | Result | DNA load (copies/<br>rxn) |  |
| **1:10<sup>2</sup>** | + | 5.2*10<sup>5</sup> | + | 3.25*10<sup>6</sup> | 16 |
| **1:10<sup>3</sup>** | + | 5.2*10<sup>4</sup> | + | 3.25*10<sup>5</sup> | 19.3 |
| **1:10<sup>4</sup>** | + | 5.2*10<sup>3</sup> | + | 3.25*10<sup>4</sup> | 22.84 |
| **1:10<sup>5</sup>** | + | 5.2*10<sup>2</sup> | + | 3.25*10<sup>3</sup> | 26.35 |
| **1:10<sup>6</sup>** | + | 5.2*10<sup>1</sup> | + | 3.25*10<sup>2</sup> | 30.80 |
| **1:10<sup>7</sup>** | - | 5.2 | + | 3.25*10<sup>1</sup> | 34 |
| **1:10<sup>8</sup>** | - | 5.2*10<sup>-1</sup> | - | 3.25 | - |

**Figure 1.**
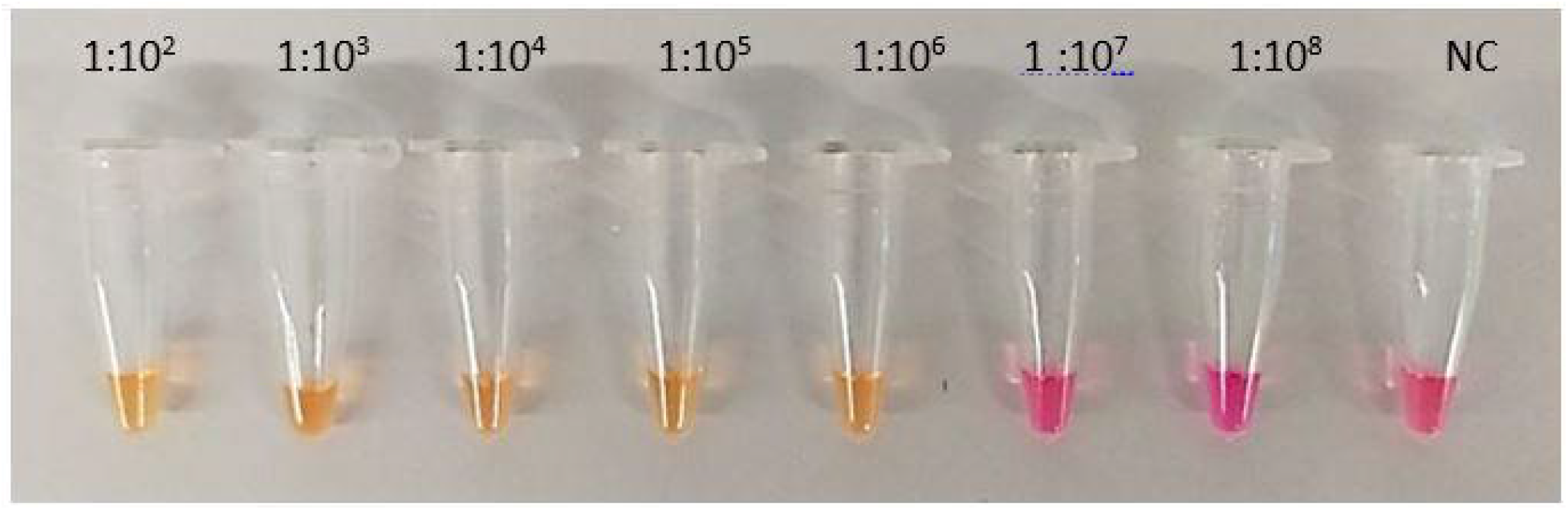
Validating the sensitivity of RT-LAMP assay. Sensitivity of RT-LAMP was determined using 1:10^2^ to 1:10^8^ serial dilutions of the N-gene which was mixed with vNAT and lysis solutions in 2:1:1 proportion, NC: Negative Control.

To validate these results, RT-PCR was performed with these dilutions (Fig.2). For RT-PCR, serial dilutions of 5*10^−3^ ng/ µL. DNA from 1:10^2^ to 1:10^9^ were prepared from the N-gene plasmid which is the same as RT-LAMP. N-gene plasmid was prepared in 1:1 proportion with vNAT and final concentrations of the diluted plasmid solution becomes 1.25*10^−2^ ng/µL. It was observed that RT-PCR can be able to detect approximately 30 copies which means nearly the same as RT-LAMP (Table 2). However, the starting volume of RT-LAMP is 25 µl and 1µl belongs to the sample. In RT-PCR, the total volume is 20 µl and 5 µl belongs to the sample. Thus, the total volume is lower, and the sample volume is higher in RT-PCR in which the copy numbers were calculated according to samples volumes per reaction. The starting DNA volume of RT-PCR is higher and more concentrated than RT-LAMP, so this result can be expected to observe.

**Figure 2.**
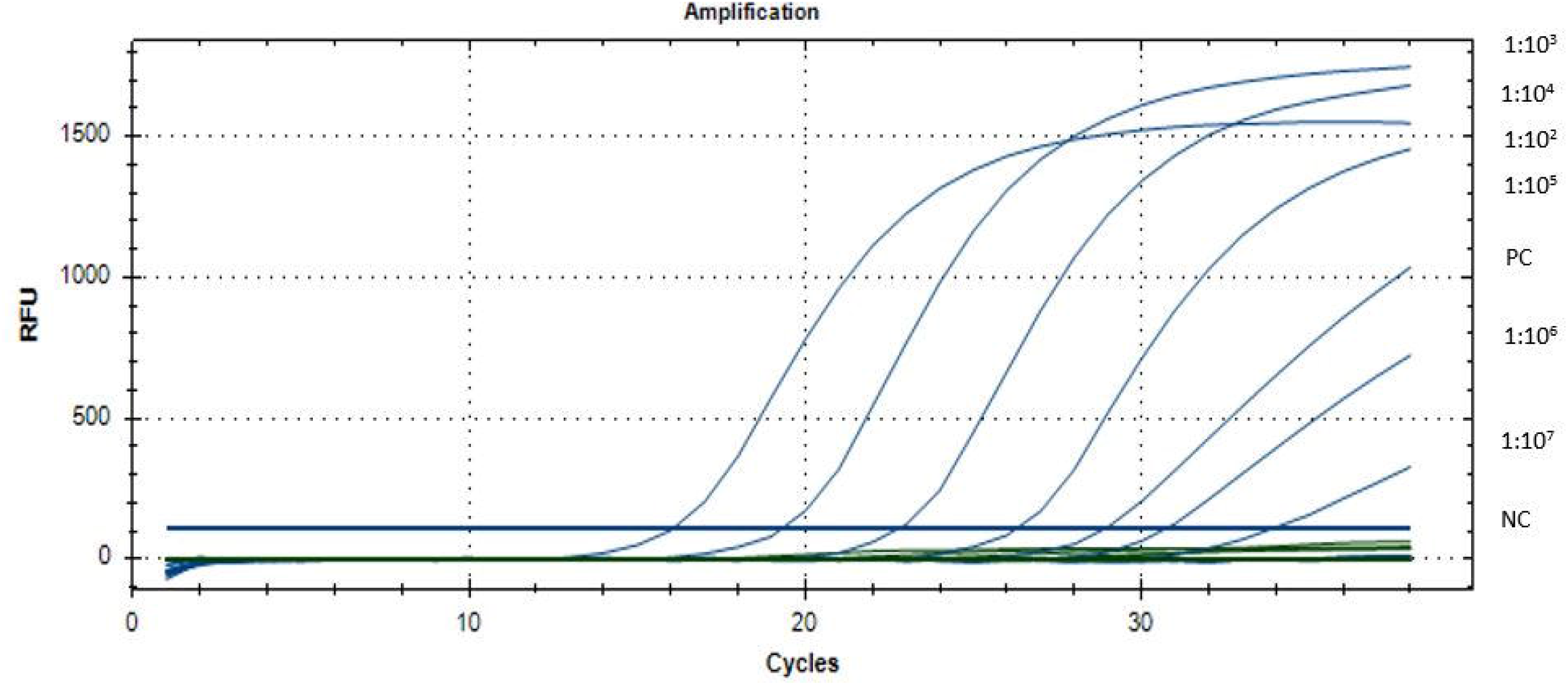
Amplification curves of RT-PCR to detect the sensitivity. Diluted N-gene in vNAT and lysis solution was used to compare the sensitivity with RT-LAMP. PC: Positive control, NC: Negative control.

### Comparison of the RT-LAMP assay and RT-PCR results with clinical samples

Analysis of the RT-LAMP and RT-PCR tests results confirmed 90 % consistency with 272 out of 300 being evaluated consistent in both the RT-LAMP and RT-PCR tests results (Table 3a). In the inconsistent group, 7 samples were tested as negative with n-Fast kit, but these samples belonged to volunteers who were diagnosed with COVID-19 with positive RT-PCR results. The remaining 21 inconsistent samples were positive with the n-Fast kit, but negative results were obtained with RT-PCR (Table 3b).

**Table 3a.** Distribution of samples according to test results.

|  | Number | Percent |
| --- | --- | --- |
| **Consistent results** | 272 | **%90** |
| **Inconsistent results** | 28 | %10 |
| **Total** | 300 | %100 |

**Table 3b.** Comparison of n-FAST kit and RT-PCR results.

| **n-Fast kit** | **RT-PCR** |  |
| --- | --- | --- |
|  | Positive | Negative |
| Positive | 24 | 21 |
| Negative | 7 | 248 |

Then, the inconsistent results were investigated more extensively by other evaluations. 21 samples were found positive with the n-Fast kit which were found negative by RT-PCR. The clinical, epidemiological, laboratory and radiological evaluations supported n-Fast kit positivity for 19 out of 21 patients. Of these patients, 7 of them have had COVID-19 recently but none of them show symptoms at the time of sample collection. So, it was concluded that the viral shedding of these patients continues. 5 of them had a close contact with a COVID-19 patient. 5 of the patients had the symptoms which are possible diagnostic criteria for COVID-19. One of the patient’s test results was positive both RT-PCR and RT-LAMP after three days from the first sample collection. Thus, RT-LAMP can detect COVID-19 at its preliminary phases. In the last case, it was determined that one patient had COVID-19 after examining his/her radiology result and it was diagnosed that the patient had lymphopenia and ground glass density in the lung compatible with early-stage COVID-19. The positivity of the remaining two n-Fast kit test results could not be supported by clinical, epidemiological, laboratory, and radiological findings (Table 4).

**Table 4.**
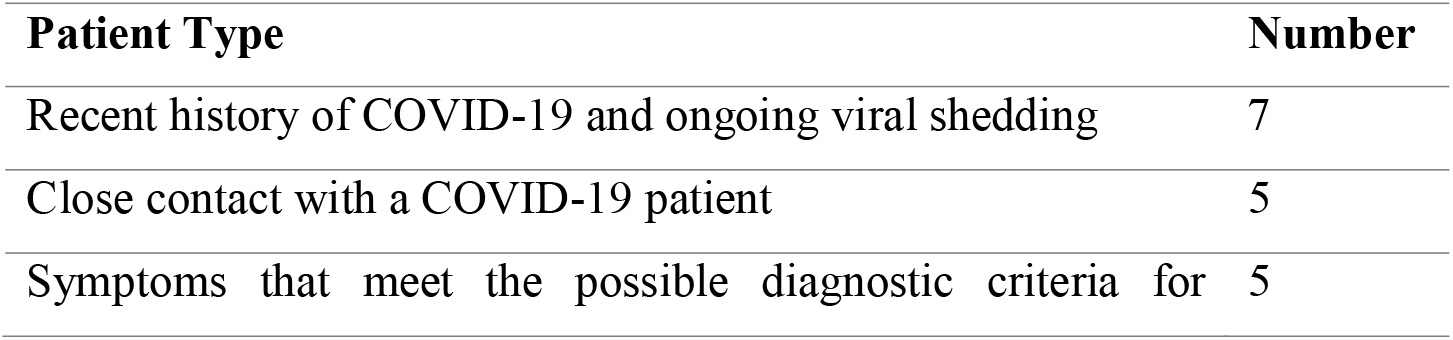
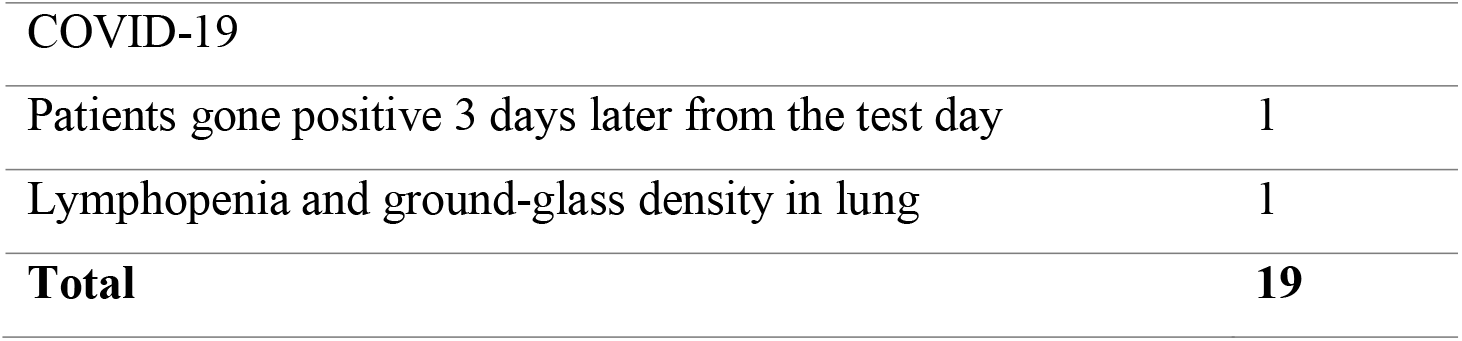
Evaluation of volunteers whose clinical, epidemiological, laboratory, and radiological findings supported n-Fast kit positivity despite negative RT-PCR results.

While the n-Fast kit results of seven volunteers were negative, they were found as positive by RT-PCR. Two of these samples were taken from asymptomatic individuals. One of these two individuals has had a medium-risk contact with a person diagnosed with COVID-19. The other has had COVID-19 recently and it was concluded that viral shedding may continue during the convalescence period. The other 5 samples were taken from patients who already showed symptoms, and the diagnoses of COVID-19 were confirmed by repeated RT-PCR results, which did not support the results of the n-Fast kit (Table 5).

**Table 5.**
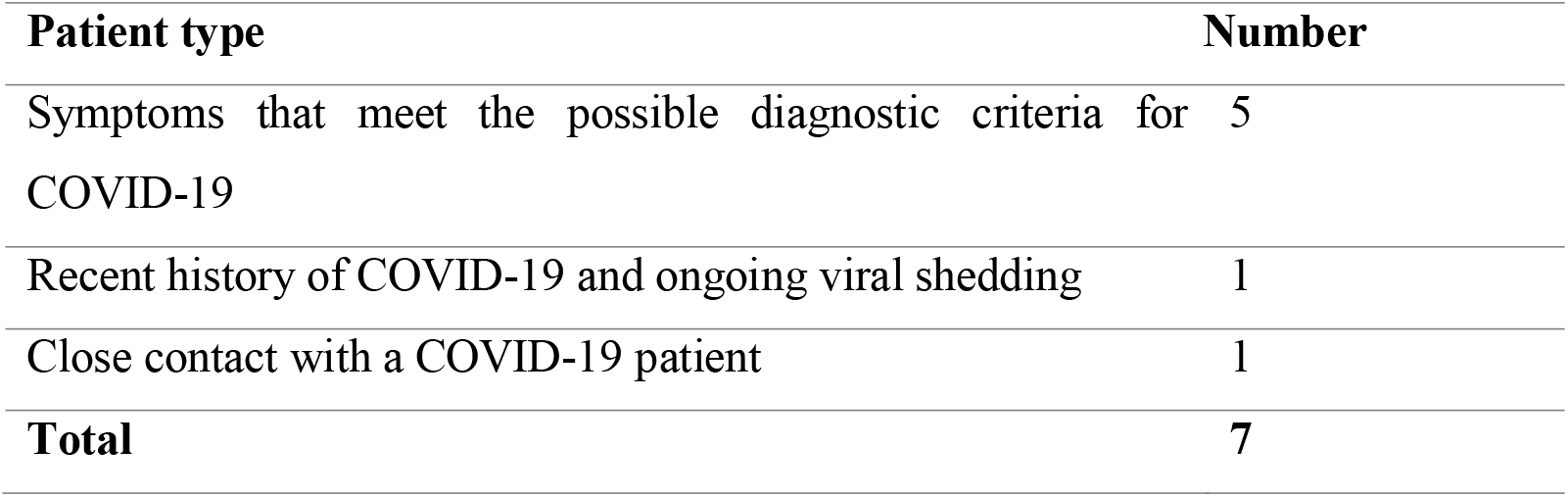
Evaluation of volunteers diagnosed with COVID-19 by RT-PCR positivity despite negative results with n-Fast kit.

| Patient type | Number |
| --- | --- |
| Symptoms that meet the possible diagnostic criteria for COVID-19 | 5 |
| Recent history of COVID-19 and ongoing viral shedding | 1 |
| Close contact with a COVID-19 patient | 1 |
| **Total** | **7** |

As shown in Table 3a, n-Fast kit test results of 272 volunteers were consistent and 28 were inconsistent with RT-PCR. In this case, the consistency between n-Fast kit and RT-PCR was found to be 90%. Being depicted in Table 4, the evaluations of the volunteers for 19 of the 28 inconsistent samples supported the results n-Fast kit. Based on these data, only 9 out of 300 volunteers were evaluated as inconsistent between n-Fast kit and RT-PCR with their clinical, epidemiological, laboratory, and radiological evaluations. Based on the final evaluation, the consistency of n-Fast kit results with RT-PCR results was calculated as 97% (Table 6). Moreover, it was calculated that the n-Fast kit has 98% specificity and 96% sensitivity according to these evaluations. Thus, the n-Fast kit is evaluated as a strong tool for the diagnosis of COVID-19 with high sensitivity and specificity.

**Table 6.** Consistency of reassessed results after clinical, epidemiological, laboratory, and radiological evaluations.

|  | Number | Percent |
| --- | --- | --- |
| **Consistent results\*** | 291 | **%97** |
| **Inconsistent results\*** | 9 | %3 |
| **Total** | 300 | %100 |

After receiving the sensitivity and specificity results of n-Fast kit, Turkish Republic Ministry of Health, Turkish Drugs and Medical Devices Agency accepted the n-Fast kit as an *in-vitro* diagnostic kit to use detection of Covid-19 in humans with the product tracking system number 8683011276071. In addition, the usage of the n-Fast kit for the diagnosis of Covid-19 is advantageous because of the detection in its early stages.

## DISCUSSION

In this study, we developed a colorimetric RT-LAMP assay for SARS-CoV-2 viral RNA and compared it to RT-PCR. N-gene plasmid serial dilution was performed to test the colorimetric RT-LAMP assay’s sensitivity and it is significantly comparable with RT-PCR. Reevaluation and further investigation of the inconsistent results demonstrated that 19 out of previously reported positive 21 cases by n-Fast kit were truly positive which greatly supports the specificity and efficiency of the n-Fast kit. Examining the inconsistencies between RT-PCR and colorimetric RT-LAMP continued with diagnostic methods like anamnesis, screening and examining radiology results. Moreover, inconsistent results between RT-PCR and colorimetric RT-LAMP were investigated with further diagnostic methods like anamnesis, screening and examining radiology results. According to these findings, 19 of the 21 inconsistent positive results were re-diagnosed as positive. In line of these results, Turkish Republic Ministry of Health, Turkish Drugs and Medical Devices Agency accepted the kit to use for Covid-19 diagnosis even in early phases of the disease. As a result, this research supports that colorimetric RT-LAMP is a cost-effective and time-saving assay and it can be considered as an accurate COVID-19 diagnosis approach.

## Data Availability

All data produced in the present work are contained in the manuscript.

## Notes

### Competing Interest Statement

The authors have declared no competing interest.

### Funding Statement

This study did not receive any funding.

### Author Declarations

Turkish Medicine and Medical Devices Agency ethics committee gave approval for 13.10.2020 dated and E.471441 numbered application.

